# The development and validation of multiplex real-time PCRs with fluorescent melting curve analysis for simultaneous detection of six bacterial pathogens of lower respiratory tract infections and antimicrobial resistance genes

**DOI:** 10.1101/2023.04.05.23288171

**Authors:** Tran Thi Ngoc Dung, Voong Vinh Phat, Chau Vinh, Nguyen Phu Huong Lan, Nguyen Luong Nha Phuong, Le Thi Quynh Ngan, Guy Thwaites, Louise Thwaites, Maia Rabaa, Nguyen Thi Kim Anh, Pham Thanh Duy

**Affiliations:** Oxford University Clinical Research Unit, Ho Chi Minh City, Vietnam; Hospital For Tropical Diseases, Ho Chi Minh City, Vietnam; Centre for Tropical Medicine and Global Health, Nuffield Department of Clinical Medicine, Oxford University, United Kingdom

**Keywords:** Real-time PCRs, lower respiratory tract infections, antimicrobial resistance, melting curve analysis, molecular diagnostics

## Abstract

*Klebsiella pneumoniae, Acinetobacter baumannii, Pseudomonas aeruginosa, Escherichia coli, Streptococcus pneumoniae and Staphylococcus aureus* are among the major bacterial causative agents of lower respiratory tract infections (LRTIs), causing substantial morbidity and mortality globally. The rapid increase of antimicrobial resistance (AMR) in these pathogens poses significant challenges for effective antibiotic therapy of LRTIs. In low-resourced settings, the diagnostics of LRTIs relies heavily on microbiological culture and patients are often treated with empirical antibiotics while awaiting several days for culture results. Rapid detection of LRTIs pathogens and AMR genes could prompt early antibiotic switching and inform antibiotic treatment duration. In this study, we developed multiplex quantitative real-time PCRs using EvaGreen dye and melting curve analysis (MCA) to rapidly identify the six major LRTIs pathogens and their AMR genes directly from the tracheal aspirate and sputum samples. The accuracy of RT-PCRs was assessed by comparing its performance against the gold standard, conventional culture method on 50 tracheal aspirate and sputum specimens. Our RT-PCR assays had 100% sensitivity for *K. pneumoniae, A. baumannii, P. aeruginosa, E. coli* and 63.6% for *S. aureus* and the specificity ranked from 87.5% to 97.6%. The kappa correlation values of all pathogens between the two methods varied from 0.63 to 0.95. The limit of detection (LOD) of target bacteria in multiplex RT-PCRs was 1600 CFU/mL. Compared to the culture results, PCR assays exhibited higher sensitivity in detecting mixed infections and *S. pneumoniae*. Our findings also demonstrated a high level of concordance between the detection of AMR gene and AMR phenotype in single infections. We conclude that our multiplex quantitative RT-PCRs with fluorescence MCA is simple but sensitive and specific in detecting six major drug resistant bacterial pathogens of LRTIs and should be further evaluated for clinical utility.

## Introduction

Lower respiratory tract infections (LRTIs) are the leading infectious cause of morbidity and mortality globally. It is estimated that 336 million episodes of LRTIs occurred in 2016, resulting in nearly 2.4 million deaths of all ages and 652,000 deaths in children less than 5 years ^1^. Although viruses such as influenza virus and respiratory syncytial virus (RSV) are responsible for a largest proportion of LRTIs, most deaths are caused by bacterial agents, including *Streptococcus pneumoniae, Haemophilus influenzae, Staphylococcus aureus, Pseudomonas aeruginosa, Klebsiella pneumoniae, Escherichia coli* and *Acinetobacter baumannii* ^2–4^.

In recent years, the worldwide increase of antimicrobial resistance (ARM) in respiratory bacterial pathogens threatens the effectiveness of antibiotic treatment ^5^. Multidrug-resistant (MDR) Gram-negative pathogens are more frequently identified from patients with LRTIs, especially in ICUs, leading to increased risks of poor outcome, prolonged hospital stay, and mortality ^6^. Early and rapid detection of LRTIs pathogens and its AMR phenotype is key to inform appropriate antibiotic therapy and reduce risks of severe complications and mortality among LRTI patients^3,7,8^.

Currently, a variety of traditional diagnostic methods are used to detect respiratory pathogens, including microscopic examination, bacterial culture, and antigen detection ^9^. These methods often require a collection of various specimens with low sensitivity and long turnaround time ^9^. Recently, several molecular PCR-based assays have been developed for the diagnostics of respiratory pathogens. Luminex assays and TaqMan array cards can detect many viruses and bacteria simultaneously ^10,11^. The BioFire FilmArray Pneumonia Panel and Pneumonia Plus Panel can detect 18 bacteria (11 Gram negative, 4 Gram positive and 3 atypical), 7 AMR markers, and 9 viruses that cause pneumonia ^12^. However, the BioFire FilmArray assays are expensive and may not be adopted for routine diagnostics, particularly in low- and middle-income (LMIC) settings ^13^.

Alternatively, multiplex real-time PCR with either fluorescent probe or fluorescent dye targeting key locally relevant bacterial-AMR gene combinations can provide a pragmatic and inexpensive diagnostic method for clinical utility. Compared to probe-based assay, dye-based assay with melting curve analysis (MCA) is less expensive and can detect more targets. The most commonly and successfully used fluorescent dye for multiplex real-time PCR is SYBR Green.

However, recently, EvaGreen (EG) dye, a third-generation, new saturating fluorescent dye are proved to be better than SYBR Green due to EG dye can be used at higher concentrations without inhibiting PCR and shows equal binding affinity for GC-rich and AT-rich regions ^14^. EG-based multiplex real-time PCR with melting curve analysis has been developed for the detection of multiple bacterial pathogens of respiratory infections ^15,16^.

Here, our study aimed to develop and validate multiplex real-time PCR with EvaGreen MCA assays (EG-mPCR assays) to detect six bacteria (4 Gram negative and 2 Gram positive) and fourteen AMR genes directly from respiratory specimens.

## Materials and methods

### Bacteria strains

To develop and validate our EG-mPCR assays, ATCC strains, clinical strains and off-target controls (*Acinetobacter lwoffii, Staphylococcus epidermidis, Shigella sonnei, Salmonella typhi, Campylobacter jejuni*) were utilized. Standard strains of *E. coli* (ATCC25922), *K. pneumoniae* (ATCC700603), *S. pneumoniae* (ATCC49619), *S. aureus* (ATCC25923), *P. aeruginosa* (ATCC27853) were purchased from the American Type Culture Collection (ATCC, Manassas, VA, USA) and subsequently cultured in Luria-Bertani (LB) agar or nutrient agar (NA). Further, eight clinical strains carrying the target AMR genes were used as positive controls for AMR gene detection assays.

### Clinical samples preparation

Tracheal aspirate (TA) or sputum samples were collected from patients admitted to Hospital of Tropical Diseases in HCMC and sent to the Microbiology Department at Hospital for Tropical Diseases for routine microbiological diagnostics. Residual samples were transferred to the Molecular Laboratory at Oxford University Clinical Research Unit (OUCRU) for the evaluation of the EG-mPCR assays.

### Microbiological Culture

The TA and the sputum samples were collected into a sterile container, followed by Gram staining and examination under direct microscopy. The samples having <10 epithelial cells and >25 leukocytes in each area upon 100x magnification were considered good quality for bacterial culture and were included in our study. Samples passing quality checking were liquefied with Sputasol liquid (Oxoid, USA) with ratio 1:1 and diluted with MRD broth (maximum recovery diluent) with ratio 1:9 before microbiological testing. Subsequently, 1μl of suspension were inoculated into blood agar, Macconkey agar and chocolate agar media (Oxoid, USA), and incubated for 24-48 hours at 35-37°C or at 35-37°C with ∼5% CO_2_. Bacterial identification and antimicrobial susceptibility testing (AST) was performed using Vitek2 automatic identification and AST system (Biomérieux, France). In TA culture, sample with bacterial growth ≥ 10^5^ CFU/mL was considered positive following the local guidelines for microbiological diagnostics.

### Extraction of nucleic acids

DNA extraction from bacterial isolates was performed using the Wizard Genomic DNA Extraction Kit (Promega, Fitchburg, USA) according to the manufacturer’s instructions. The quality and concentration of the DNA was assessed using a Nano-drop spectrophotometer prior to PCR amplification.

For TA/sputum samples, an aliquot of 200μl was homogenized with 4 times the volume of 0.1% dithiothreitol (DTT) for 15 minutes at room temperature and centrifuged at 8000 rpm for 10 minutes. Subsequently, supernatant was discarded and pellet was resuspended with 200μl PBS. Next, 25μl of 10X buffer (200mM Tris-HCl, 0.2μl of Benzonase (Sigma) and 24.8μl of sterilized water) were added, followed by 2-hour incubation at 37°C. The mixture was centrifuged at 8000 rpm for 10 minutes and the pellet was subsequently resuspended in a mixture of 4μl EDTA, 120μl NaCl and 76μl H_2_O. Centrifugation was performed again at 8000 rpm for 10 minutes and pellet was resuspended in 200μl of TE before DNA extraction. 200μl of pre-treated samples was subjected to an automated extraction on a MagNA Pure 96 nucleic extraction system (Roche applied sciences, UK), according to the manufacturers recommendations.

### Primer Design

Twenty primer sets were designed based on the conserved regions which can produce amplicons having melting temperatures (*Tm*s) ranging from 75°C to 92°C and target-specific *Tm* values differed from each other by at least 1°C. The primer sequences and their optimal concentrations are described in Table 1. The actual *Tm* of all primers was determined following the performance of singleplex PCRs and multiplex PCRs.

**Table 1.**
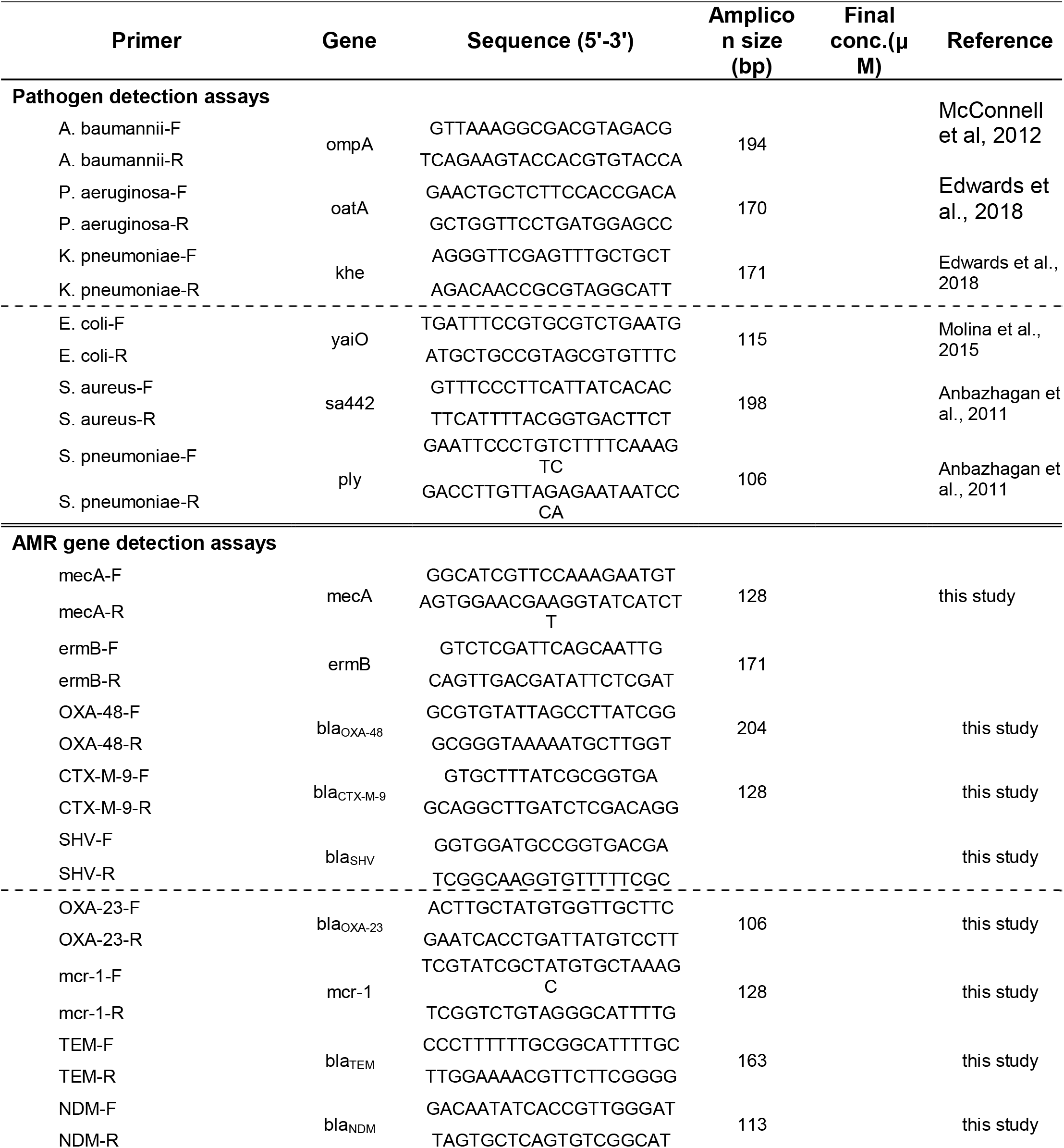
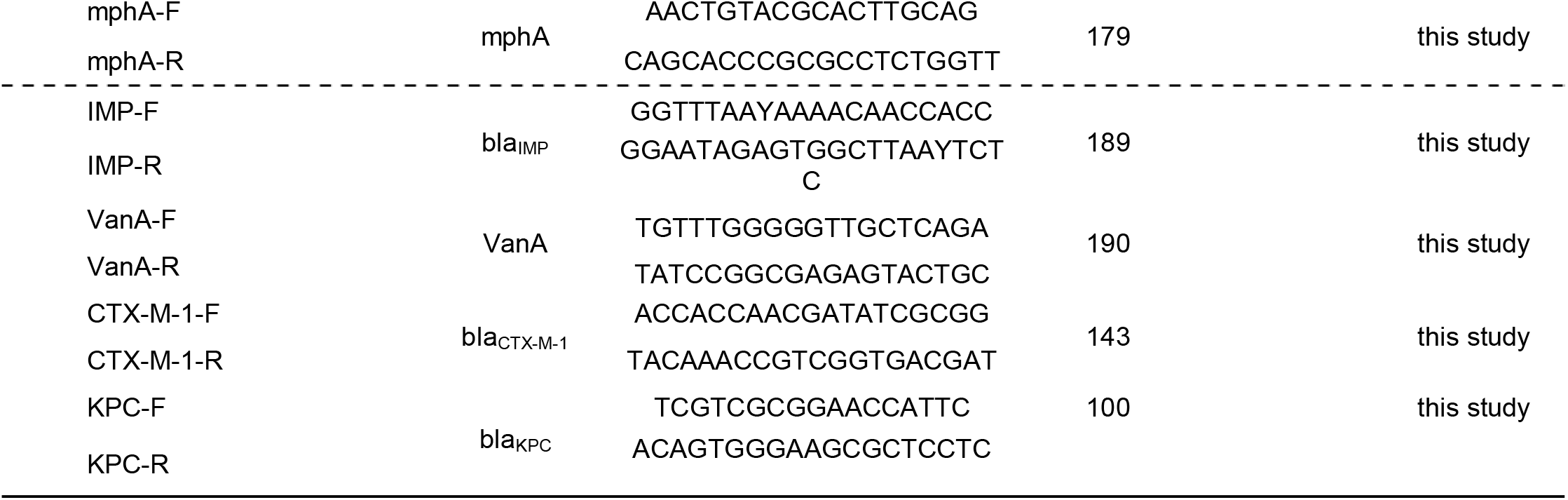
Sequences, concentrations of primers used for singleplex and multiplex real-time PCR assays

For simultaneous detection of the six LRTIs pathogens, six primer pairs were designed targeting the known species-specific genes that have been published previously (*yaiO* for *E. coli* ^17^; *ompA* for *A. baumannii* ^18^; *oatA* for *P. aeruginosa* ^19^; *sa442* for *S. aureus* ^16^; *ply* for *S. pneumoniae* ^16^ and *khe* for *K. pneumoniae* ^19^. Additionally, fourteen primers were also designed to identify fourteen common acquired AMR genes present in these bacteria, encoding resistance to β-lactams (*bla*_SHV_, *bla*_TEM_, *bla*_CTX-M-9,_ *bla*_CTX-M-1_), methicillin (*mecA*), carbapenems (*bla*_KPC_, *bla*_NDM_, *bla*_OXA-48_, *bla*_OXA-23,_ *bla*_*IMP*_), colistin (*mcr-1*), macrolides (*ermB, mphA*) and vancomycin (*vanA*).

### Development of EG-mPCR assays

Singleplex real-time PCR assays were performed in 20μl reaction volume, which included 5μl DNA template, 0.8μl forward and reverse primers and 10μl SensiFAST HRM kit (Meridian Bioscience), to check the actual *Tm*s of each individual target amplicon. Sterile purified water was used as the negative control. PCR amplifications were run on a LightCycler 480II (Roche applied sciences, UK) with the following thermal conditions: initial denaturation at 95°C for 5min, followed by 40 cycles of denaturation, 95°C, for 10 s; annealing, 60°C for 30 s; extension, 72°C for 30s. For multiplex real-time PCR assays, the component and thermal condition were the same as singleplex real-time PCRs, except the concentration of each primer pair varied from 0.25 μM to 0.5 μM. After PCR amplification, the melting curve analysis (MCA) was conducted in the same thermocycler at 65°C to 95°C; and cooling cycle at 37°C for 30 s. Fluorescence was continuously measured and the melting temperature (*Tm*) was calculated by plotting the negative derivative of fluorescence over temperature versus temperature (−d*F*/d*T* versus *T*). Conventional PCRs with a different set of primers were later performed to confirm the results of EG-mPCR assays.

The reproducibility, linearity, limit of detection (LOD) of EG-mPCR assays for pathogen detection were evaluated using 5-fold serial dilutions of two DNA control mixes. Each DNA control mix was prepared by pooling equal volume of 10^8^ CFU/mL of each bacterium, followed by DNA extraction. Subsequently, 5-fold serially diluted DNA standards, corresponding to the bacterial concentrations from 10^6^ to 12.8 CFU/mL, were prepared for the assays.

To validate the assays in TA specimen, 200μl pre-treated culture-negative TA specimen was spiked with 5-fold serial dilutions of bacteria (from 10^6^ to 12.8 CFU/mL), followed by DNA extraction using Roche’s MagNA Pure 96 system. The EG-mPCR assays were performed as described above.

### Diagnostic performance of EG-mPCR assays in comparison to conventional culture

Fifty TA and sputum samples were diagnosed using both the EG-mPCR assays and conventional microbiological culture and the results were compared between the two methods. The samples that were culture-negative but EG-mPCR positive were further confirmed by singleplex EG-PCR and conventional PCR. In addition, the correlation between the presence of AMR gene and AMR phenotype was also examined. The degree of agreement between EG-mPCR assays and conventional culture for bacterial identification was measured by Cohen’s kappa ^20^.

### Ethic statement

The study did not collect patient’s information or require taking additional samples from patients. Informed consent is waved by the Institutional Ethical Review Committee of Hospital of Tropical Diseases in Ho Chi Minh city.

## RESULTS

### Detection of six bacteria and AMR genes by EG-mPCR assays

Two EG-mPCR assays were used to detect 6 bacterial pathogens and additional three EG-mPCR assays were used to detect 14 AMR genes (Table 1). In each EG-mPCR assay, the melting curve analysis showed distinct Tm peaks corresponding to the target bacteria and AMR genes, while negative controls did not show any signal (Figure 1). The difference between two consecutive peaks was more than 2°C (Table 2). There was no interference and cross-reactivity in our EG-mPCR assays. We additionally evaluated our assays on other closely related bacteria and did not find any non-specific signal for *Salmonella* Typhi, *Campylobacter jejuni, Acinetobacter lwoffii, Staphylococcus epidermidis*. However, the *yaiO* primer which was supposed to be specific to *E. coli*, gave false positive signal for one *Shigella sonnei* isolate.

**Table 2.**
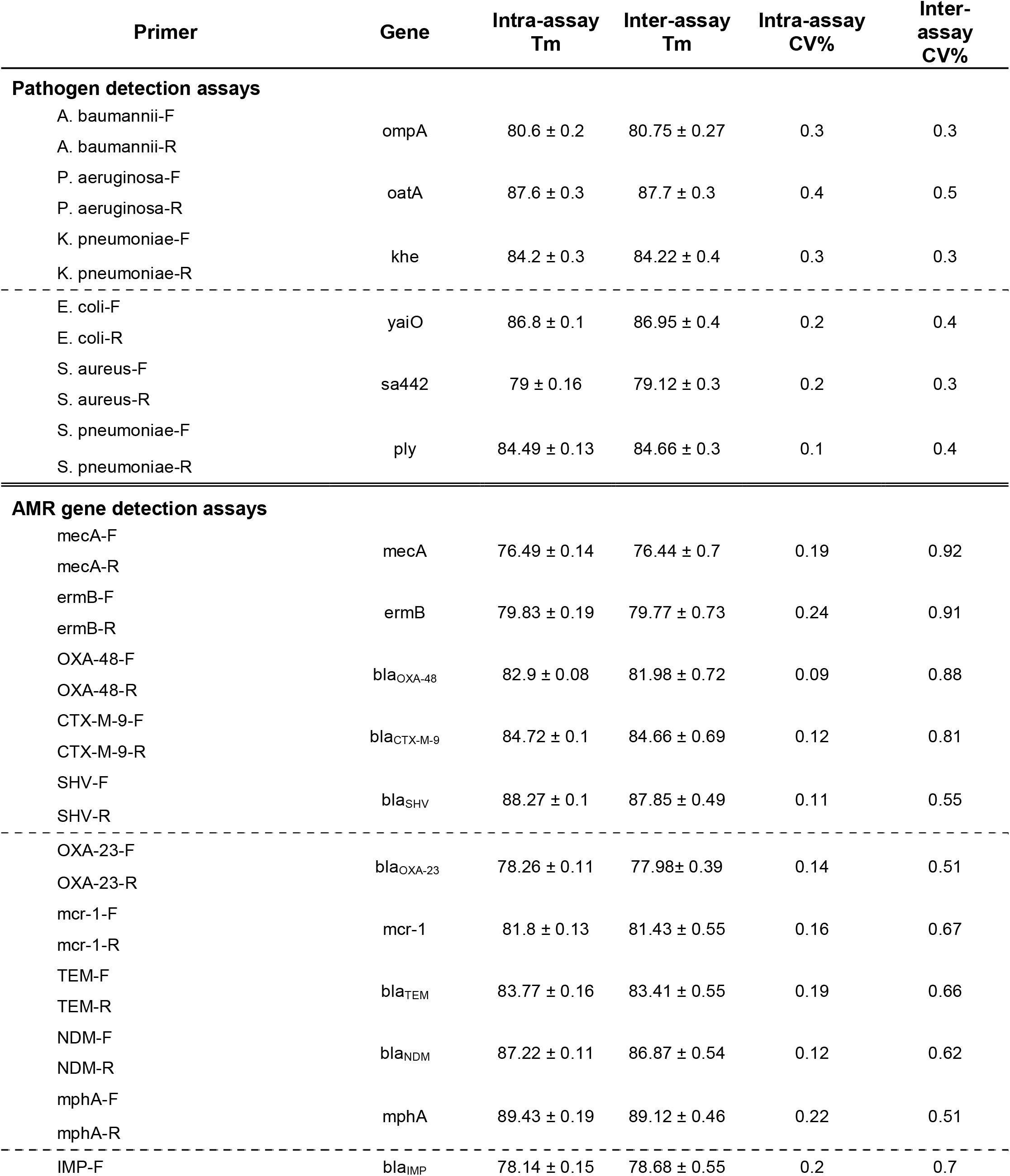
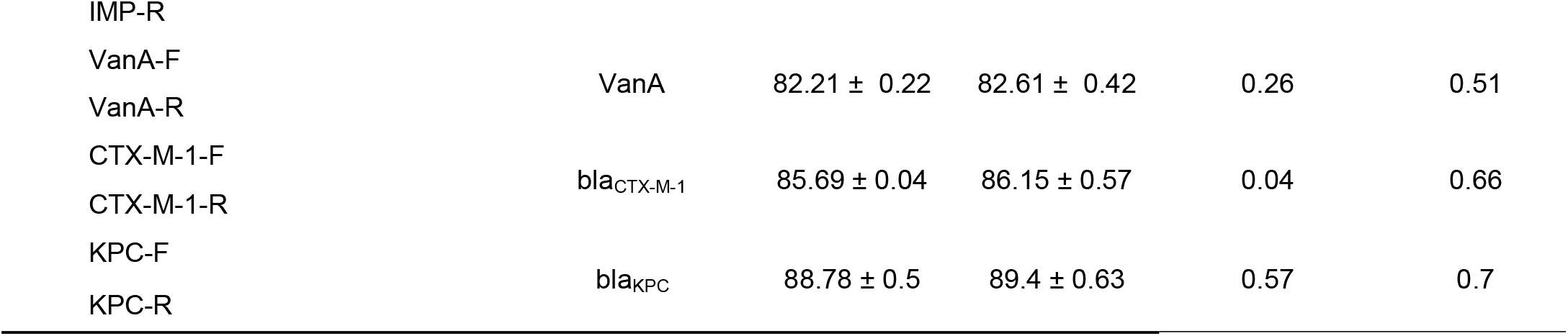
Tm values of primers used for singleplex and multiplex real-time PCR assays

**Figure 1.**
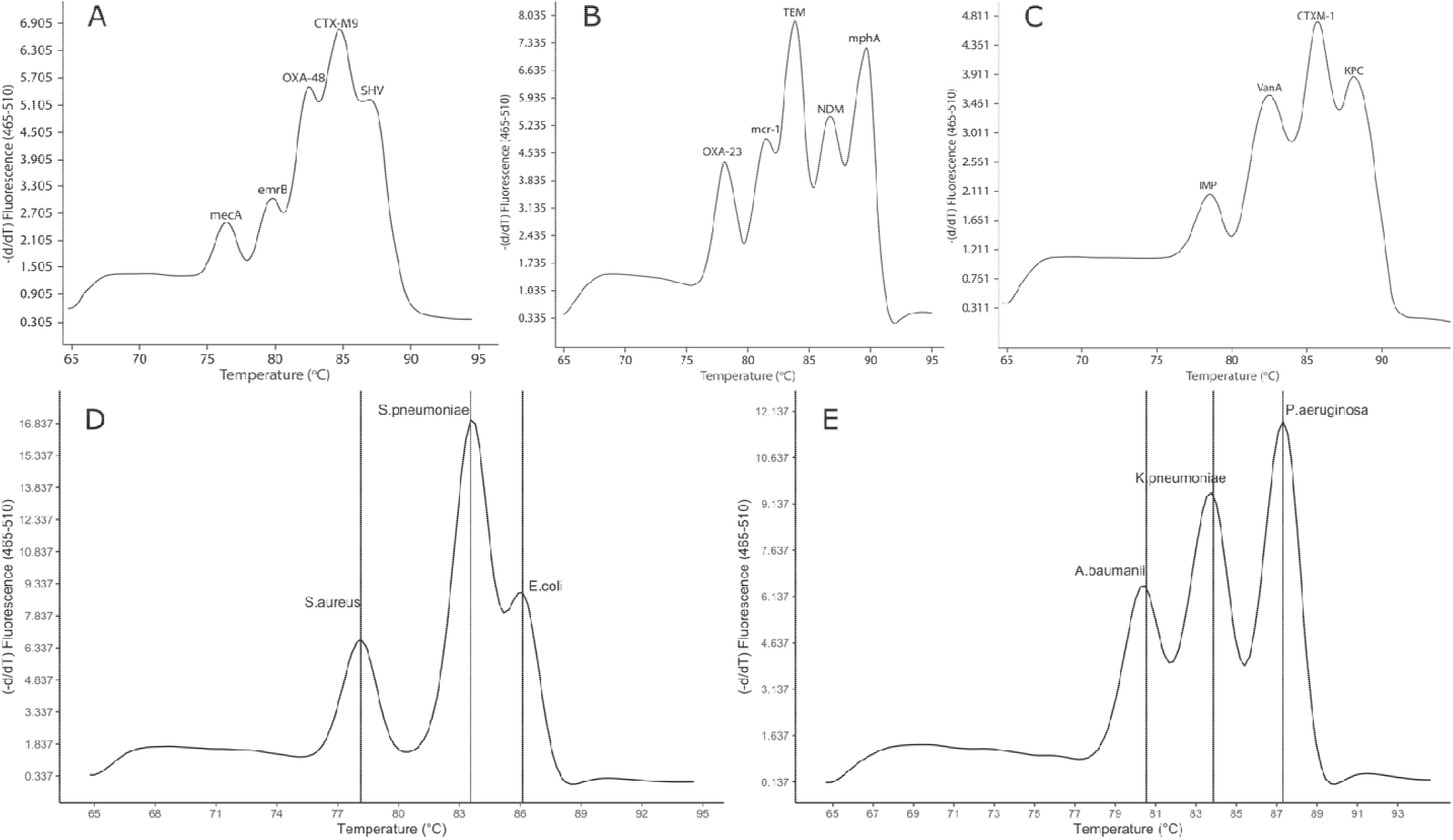
Melting curve analysis showing the melting temperature peaks (Tm) of 14 AMR genes (A, B, C) and 6 bacterial pathogens (D, E) and negative control (NTC) in 5 multiplex PCRs.

### Reproducibility, quantification and LOD of the two EG-mPCR assays for bacterial identification

The reproducibility of EG-mPCR assays was evaluated by accessing *Tm* values in inter and intra-assays. The standard deviation (SD) and coefficient variation (CV) of *Tm* were calculated in 5 replicates within the same run (intra-assay) and across 5 different runs (inter-assay). The intra-assay CV of each of the target bacteria ranged from 0.1% to 0.4% and the inter-assay CV ranged from 0.3% to 0.5%. Similarly, intra-assay CV and inter-assay CV of each of the target AMR genes varied from 0.04% to 0.57% and from 0.51% to 0.92%, respectively (Table 2). These data indicated good reproducibility of Tm values in our assays.

The standard curve for each bacterium in the two EG-mPCR assays was obtained by plotting the fluorescence intensity (−d*F*/d*T*) or the height of the peak (*y*-axis) values against the log_10_ of the bacterial concentrations (*x*-axis) inferred from the serially diluted DNA control mixes. The coefficient of determination of linear regression model of standard curves were R^2^=0.92 for *S. aureus*, R^2^=0.92 for *S. pneumoniae, R*^2^=0.91 for *E. coli*, R^2^=0.97 for *A. baumannii*, R^2^=0.98 for *K. pneumoniae* and R^2^=0.95 for *P. aeruginosa*. These data indicated a good linear correlation between the fluorescence values and the log_10_ of bacterial concentration over a range of bacterial concentrations.

The limit of detection (LOD) was evaluated by observing the Tm peaks of each bacterium in the two serially diluted DNA control mixes. The experiment was performed in 5 replicates within the same run and in 10 replicates between different runs. Based on the lowest concentration of a target bacterium in a control mix where all targets showed positive signal, a LOD of 1600 CFU/mL was identified for each of the two EG-mPCR assays (Figure 2). When a TA specimen was spiked with target bacteria, a LOD of 3200 CFU/mL was identified for each of the two EG-mPCR assays.

**Figure 2.**
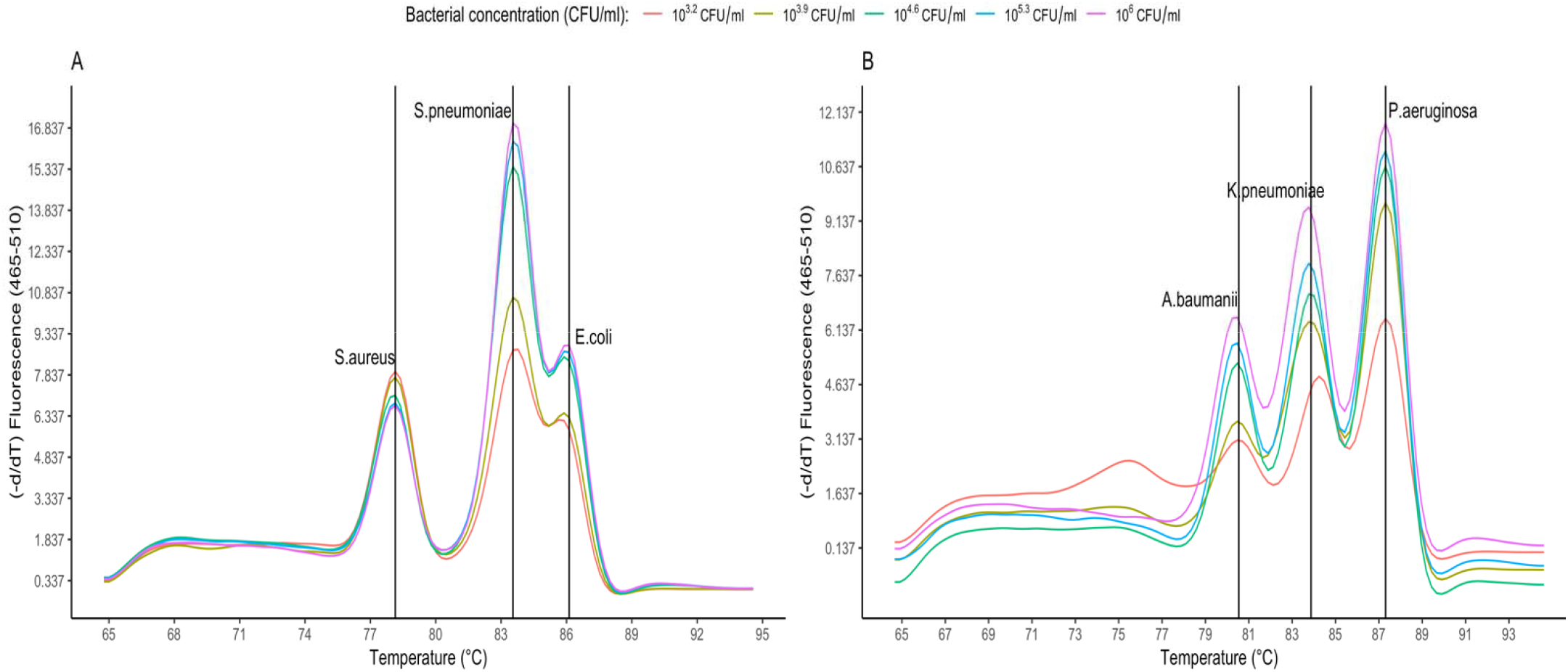
The sensitivity of simultaneous detection of *S. aureus, S. pneumoniae, E. coli* (A) and *A. baumannii, K. pneumoniae, P. aeruginosa* (B) with concentrations ranging 10^6^ to 12.8 CFU/ml. LOD of simultaneous detection of DNA of *S. aureus, S. pneumoniae, E. coli* was 1600 CFU/ml and the same for *A. baumannii, K. pneumoniae, P. aeruginosa*.

### Diagnostic performance of EG-mPCR assays in comparison to conventional culture

Between December 2020 and March 2021, 50 consecutive TA and sputum specimens were included for both conventional culture and EG-mPCR assays. Conventional culture showed that 76% (38/50) of samples were culture positive for single target pathogen and 16% (8/50) were culture positive for more than one pathogen (including 4 pathogens not covered in the PCR detection assays: *Haemophilus influenzae, Moraxella, Stenotrophomonas maltophilia, Enterobacter*). *P. aeruginosa* was the most prevalent pathogen (26%, 13/50), followed by *A. baumannii* (20%, 10/50), *S. aureus* (18%, 9/50), *K. pneumoniae* (12%, 6/50) and *E. coli* (2%, 1/50) (Table 4). Three samples (6%) were culture negative for all six pathogens. Overall, the microbiological culture from TA and sputum samples yielded a total of 51 target pathogens.

**Table 3.**
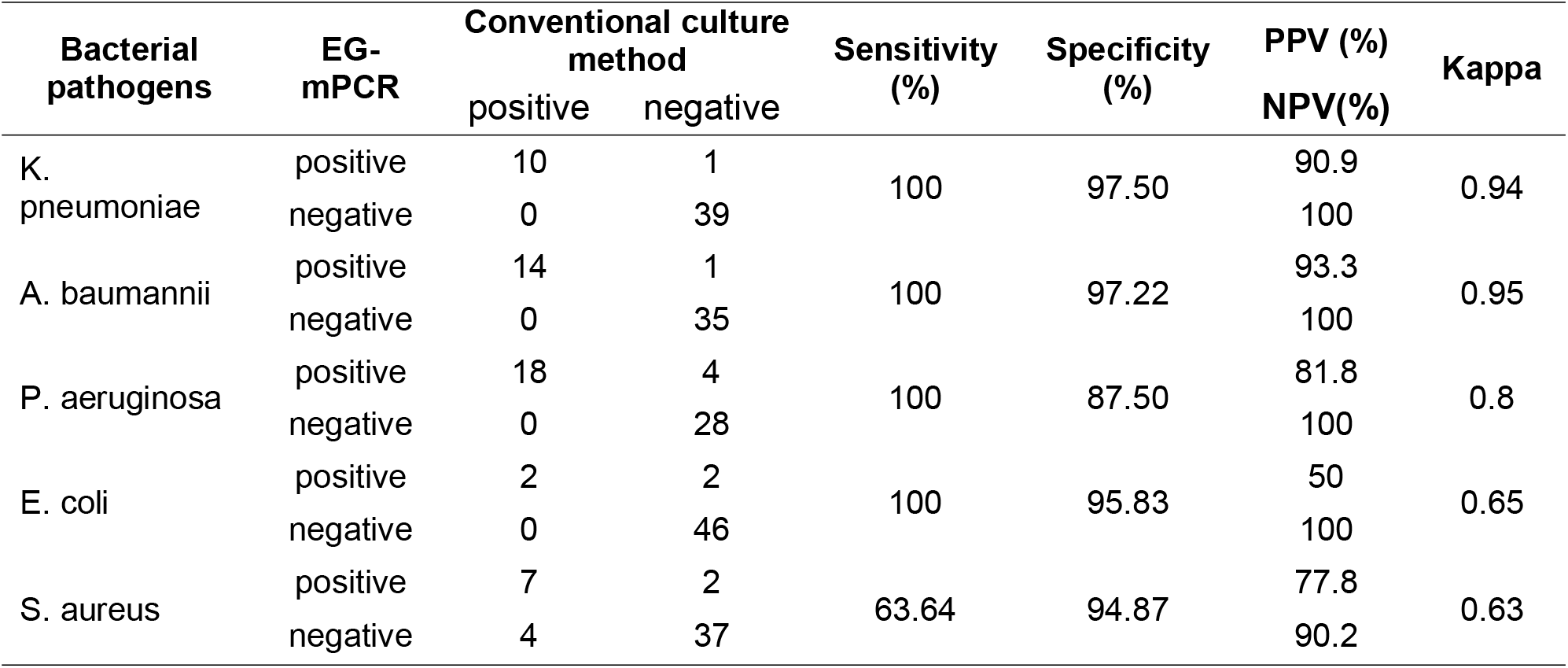
Performance of EG-mPCR assays compared with the conventional culture method

**Table 4.** Distribution of AMR genes in single infections identified by PCR assays

|  |  | Cephalosporin resistance |  | Carbapenem resistance |  | Macrolide resistance |  |
| --- | --- | --- | --- | --- | --- | --- | --- |
| Pathogens detected by culture (n) |  | n | resistant genes | n | resistant genes | n | resistant genes |
| Gram negative | *A. baumannii* (9) | 7 | TEM (7/7) | 7 | OXA23 (7/7) |  |  |
|  | *K. pneumoniae* (5) | 4 | TEM (3/4) | 2 | NDM (2/2) |  |  |
|  |  |  | CTXM-1 (1/4) |  | OXA-48 (2/2) |  |  |
|  | *E. coli* (2) | 1 | SHV (1/1) | 1 |  |  |  |
|  |  |  | TEM (1/1) |  |  |  |  |
|  | *P. aeruginosa* (9) | 1 | CTXM-9 (1/1) | 1 | OXA23 (1/1) |  |  |
| Gram positive | *S. aureus* (5)* |  |  |  |  | 5 | emrB (3/5) |

\* EG-mPCR detected 3 *S. aureus* and did not identify 2 *S. aureus* in single-infection cases.

According EG-mPCR results, 56% (28/50) of samples had one peak with strong fluorescence signal and were identified as single infections. There were 18 samples (36%) showed at least 2 melting curve peaks with strong fluorescence signals, which were identified as co-infections.

The most common patterns of co-infections detected by PCR were *P. aeruginosa* and *S. pneumoniae* (3/18) and *P. aeruginosa* and *S. aureus* (3/18), *P. aeruginosa* and *K. pneumoniae* (2/18) and *P. aeruginosa* and *A. baumannii* (2/18). *P. aeruginosa* (12/18) was the most common pathogen detected in co-infections (Figure 3). EG-mPCR showed negative results in 4 samples, three of which were also culture negative. Overall, EG-mPCR assays yielded a total of 67 target pathogens.

**Figure 3.**
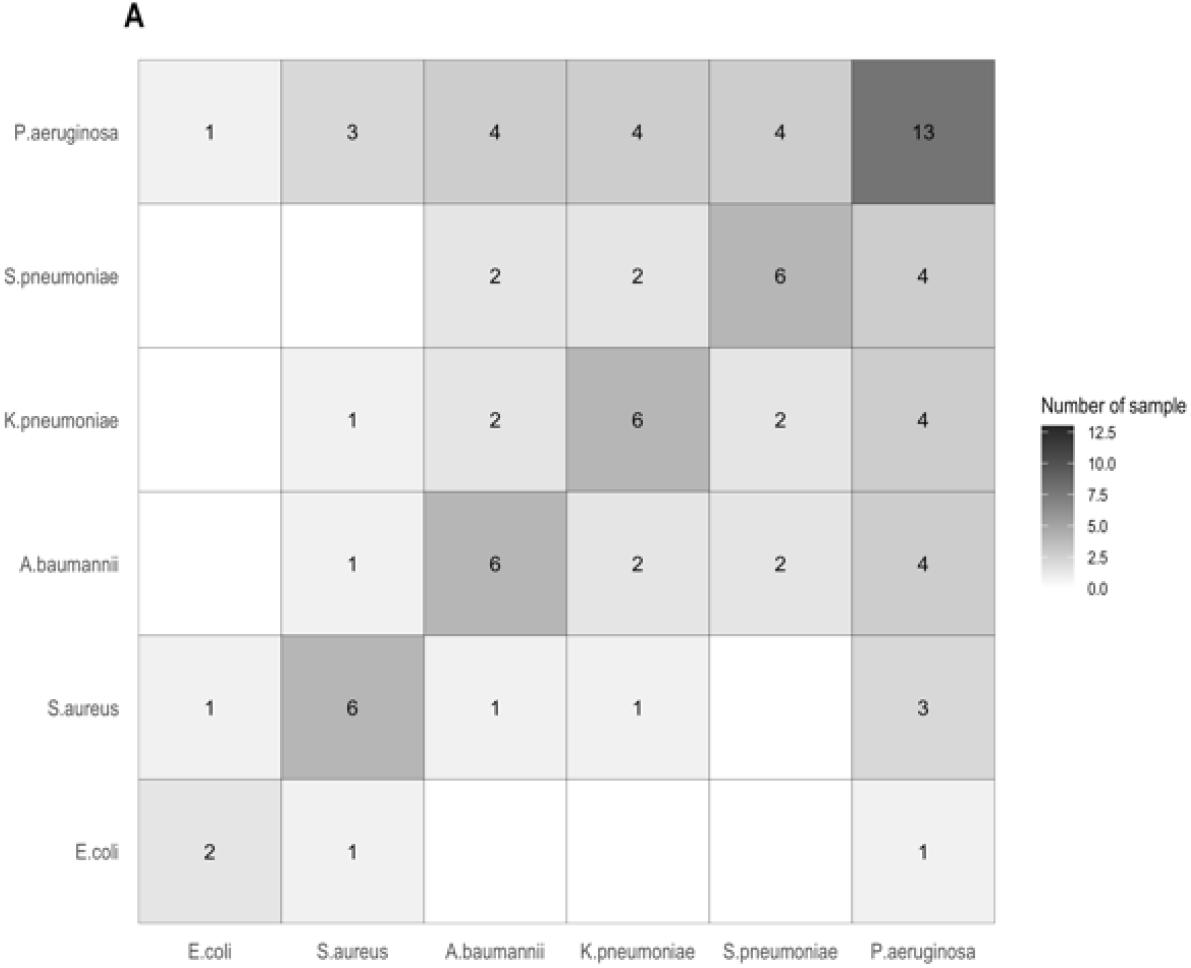
Heatmap of bacterial mixed infection pattern detected by the EG-mPCR method in the clinical samples.

Compared to conventional culture, the sensitivity and specificity of EG-mPCR assays for *K. pneumoniae, A. baumannii, P. aeruginosa, E. coli, S. aureus* were 100% and 97.5%, 100% and 97.2%, 100% and 87.5%, 100% and 93.8%, 63.6% and 94.9%, respectively (Table 3). The positive predictive value (PPV) and negative predictive value (NPV) of EG-mPCR assays were 90.9% and 100% for *K. pneumoniae*, 93.3% and 100% for *A. baumannii*, 81.8% and 100% for *P. aeruginosa*, 50% and 100% for *E. coli* and 77.8% and 90.2% for *S. aureus*, respectively.

Out of the microbiologically-confirmed 51 target pathogens, 48 pathogens (94.1%) were also detected by EG-mPCR assays with a quantitative result ≥ 10^5^ CFU/mL, which is a cut off of culture positivity (Figure 4A). Our data demonstrated a high degree of similarity between EG-mPCR assays and culture method. When considering only microbiologically-confirmed target pathogens with growth ≥ 10^5^ CFU/mL (n=26) in TA samples, concordant PCR-based quantitative results were found in 100% (11/11) of *A. baumannii*, 100% (1/1) of *E. coli*, 100% (5/5) of *K. pneumoniae*, 100% (8/8) of *P. aeruginosa* and 100% (1/1) of *S. aureus* (Figure 4B). There were however discordant results between the two methods. The culture method did not grow any *S. pneumoniae* isolates whereas the PCR assay found 6 *S. pneumoniae* with a quantity ranging from 10^3.6^ to 10^9^ CFU/mL; furthermore, the culture failed to detect another 10 target bacteria that were identified by PCR (8 bacteria with a quantity ≥ 10^5^ CFU/mL and 2 bacteria with a quantity <10^5^ CFU/mL) (Figure 4A). Consequently, a total of 26 isolates were identified by PCR as mixed infections, for which only 11 isolates were identified by culture method as single infections and 15 isolates were missing from the culture results.

**Figure 4.**
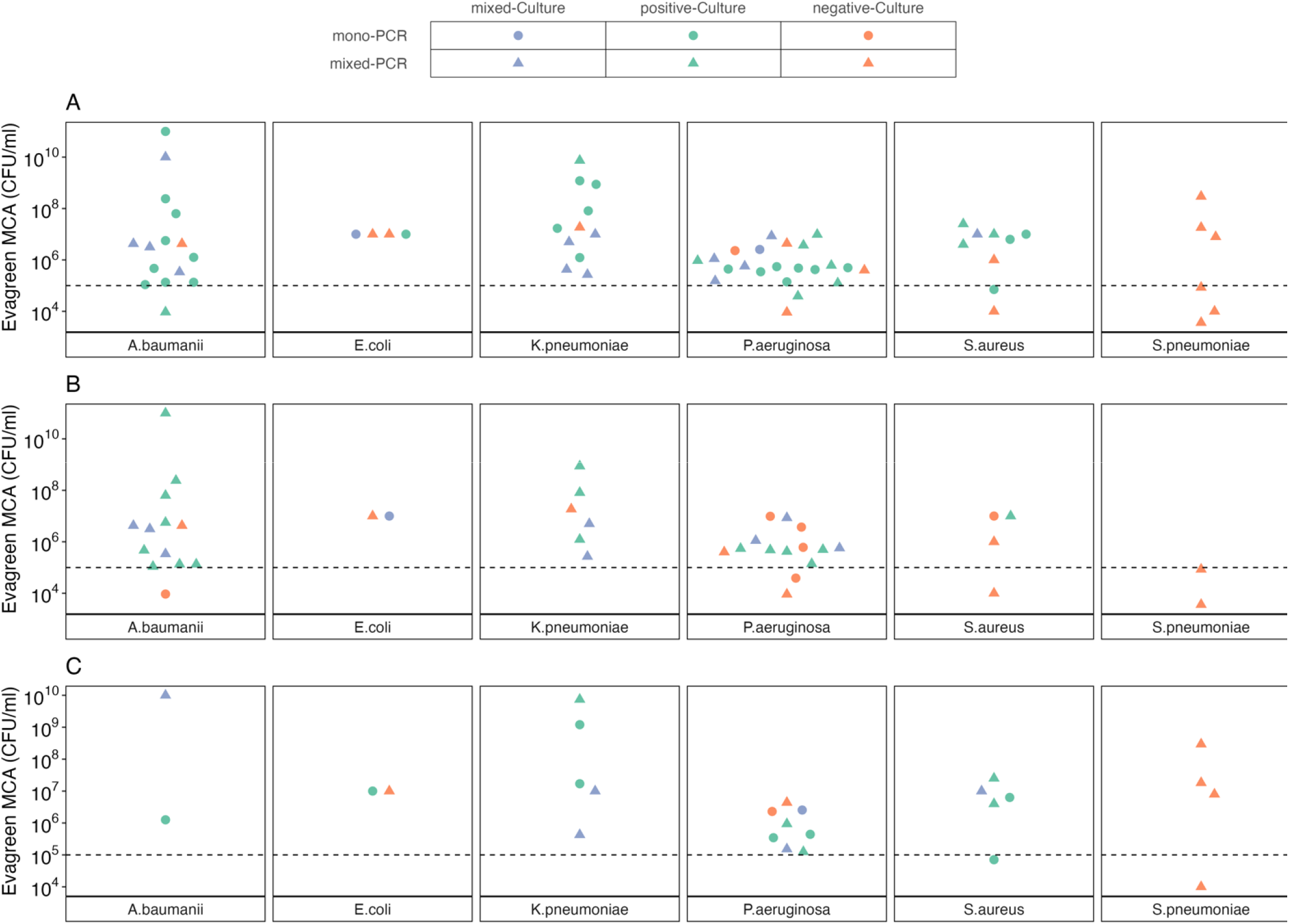
The detection and quantity of 6 target bacteria by EG-mPCR assays in accordance with culture results (culture-negative, culture-positive with one target bacterium, culture-positive with at least two target bacteria)

**Figure 5.**
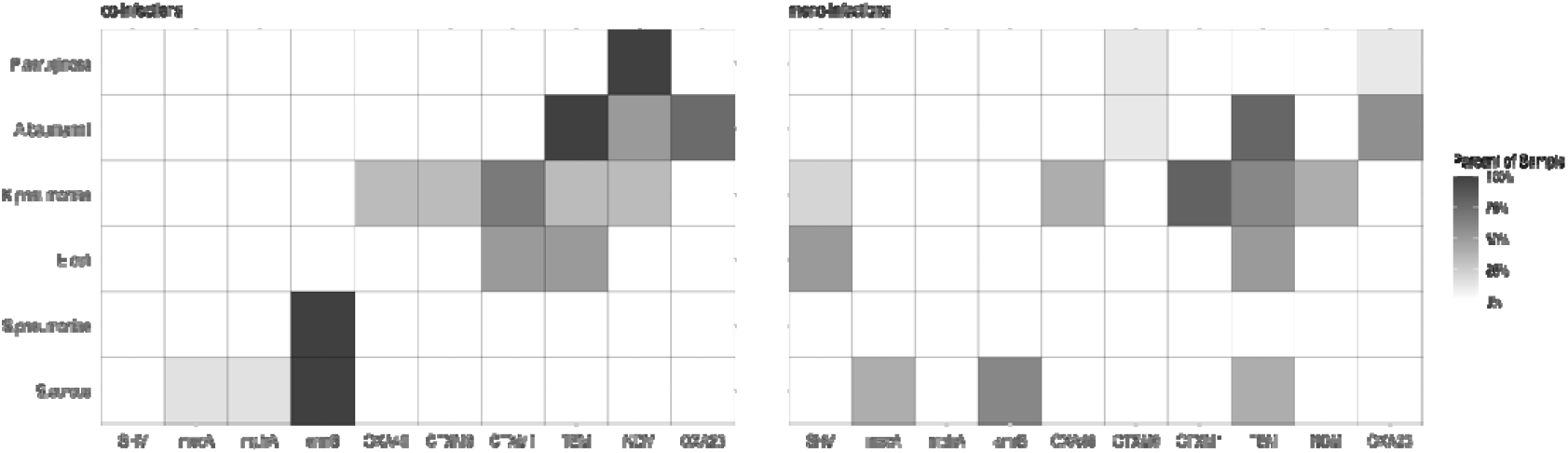
The distribution of predicted AMR genes in co-infections

### Application of EG-mPCR assays for AMR gene detection from clinical samples

Three EG-mPCR assays were used to identify 14 AMR genes commonly present in the target pathogens from 50 clinical samples. After excluding mixed infections by PCR, the level of correlation between the presence of AMR gene and AMR phenotype was assessed in 28 target bacteria. The overall distribution of AMR genes and AMR profiles of 28 target bacteria are shown in Table 4 and Figure 6. In *A. baumannii*, 100% (7/7) of the isolates carried *bla*_OXA23_ and *bla*_TEM_ were resistant to ceftazidime, cefepime and carbapenem (imipenem, meropenem).

**Figure 6.**
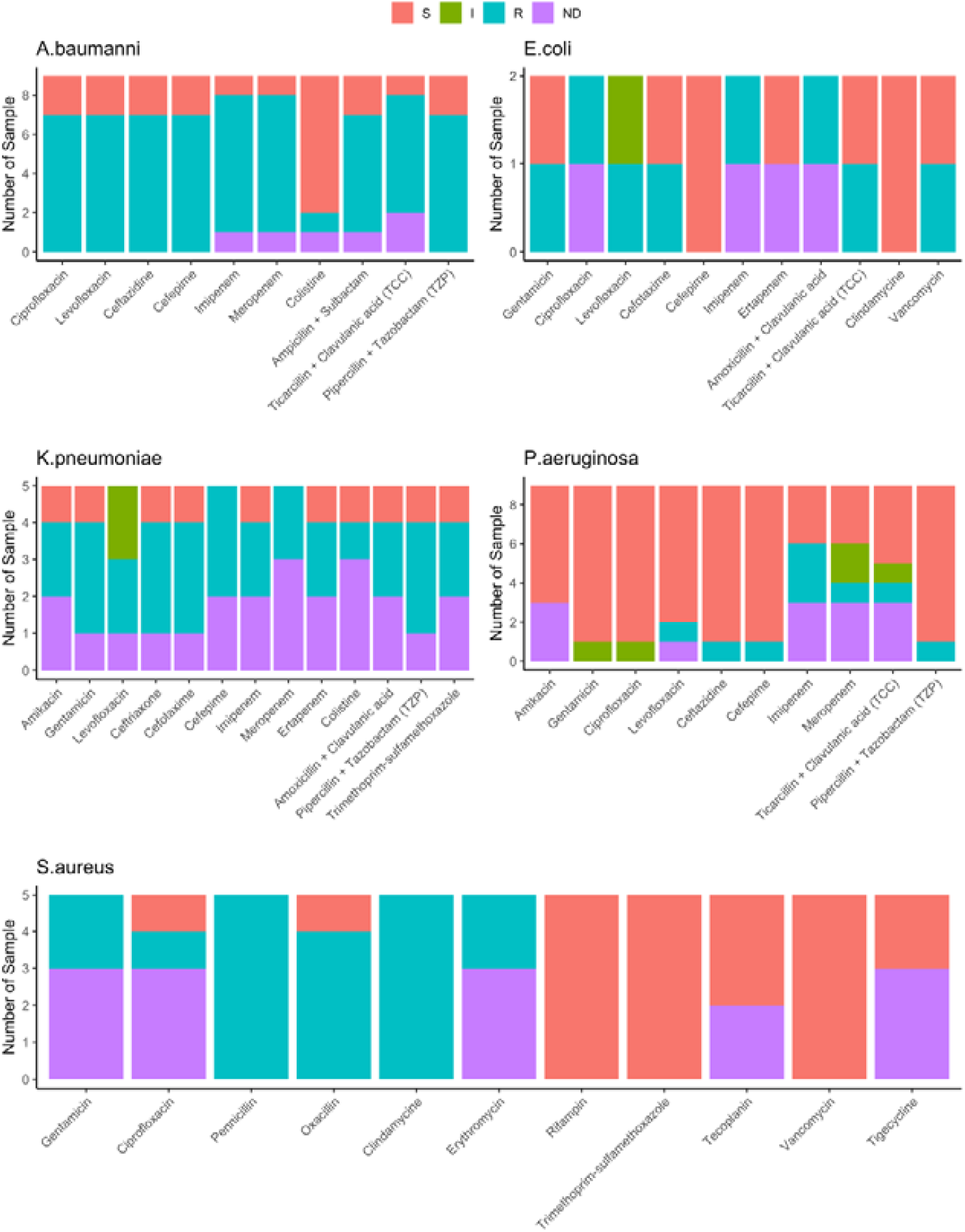
The antimicrobial susceptibility testing results of target bacteria.

Among 4 *K. pneumoniae* isolates that were resistant to 3^rd^ and 4^th^-generation cephalosporins (ceftriaxone, cefotaxime, cefepime), *bla*_CTX-M-1_ was found in all 4 isolates and *bla*_TEM_ was found in 3/4 isolates. *Bla*_NDM_ and *bla*_OXA48_ genes were both detected in two carbapenem resistant *K. pneumoniae* isolates. For *P. aeruginosa, bla*_OXA23_ was found in one isolate that was resistant to imipenem and meropenem and *bla*_CTX-M-9_ was found in another isolate that was resistant to the 3^rd^ generation cephalosporin (ceftriaxone, cefotaxime). Among five *S. aureus* isolates that were resistant to macrolides (clindamycin), *emrB* gene was detected in 3 isolates (60%). Two isolates (1 *K. pneumoniae* and 1 A. *baumannii*) were resistant to colistin, but no resistance genes were found. There was one *E. coli* isolate resistant to cefotaxime and imipenem for which *bla*_SHV_ and *bla*_TEM_ were identified but carbapenem resistance genes were undetected.

## DISCUSSION

The selection and duration of effective empirical antibiotic treatment for LRTIs have become a major challenge in LMIC settings where MDR causative bacteria are prevalent. Although culture-guided definitive treatment is often followed to avoid overuse/misuse of initial antibiotics, the long waiting time for culture results has delayed effective antibiotic therapy, posing increased risks of mortality and selection of antimicrobial resistant organisms. Rapid and inexpensive molecular PCR-based diagnostics is urgently needed in clinical practice to inform antibiotic therapy. Here, we successfully developed two EG-mPCR assays to detect and quantify 6 major bacterial pathogens and three EG-mPCR assays to identify 14 AMR genes directly from TA and sputum samples. The two PCR assays for pathogen detection exhibited high sensitivity (from 63.6% to 100%) and high specificity (from 87.5% to 97.5%) compared to conventional culture. The turnaround time from sample collection to the final PCR results was less than 6 hours.

A key advantage of our PCR assays is the ability to quantify the amount of each bacterium in a multiplex PCR reaction and thus could distinguish between bacterial colonization and infection. In fact, the quantitative data from the PCR assays demonstrated 100% concordance with quantitative microbiological culture results of TA samples. A previous study using a similar method raised concern about the reduced linear relationship between fluorescence levels and bacterial concentration in multiplex PCR assays, which may be attributed to the competition amongst EvaGreen dyes ^15^. This limitation has been addressed in this work by increasing the amount of the PCR mix and adding extra EvaGreen dye to the reaction. Our experiments showed a good linear correlation (R^2^ >0.91) between fluorescence readings and the log10 of bacterial concentrations. Furthermore, the LOD of PCR assays (10^3.6^ CFU/mL) was much lower than the cut-off points for culture positivity (≥10^5^ CFU/mL), meaning our assays not only could identify true pathogens above the cut-off but also detect other potentially pathogenic bacteria present with lower concentrations for further monitoring.

Notably, PCR assays were able to detect more mixed target bacteria in the respiratory samples. The facts that EG-mPCR assays were more sensitive than conventional culture in detecting mixed infections, of which most target bacteria had a concentration >10^5^ CFU/mL suggested that culture method may have missed some true target pathogens in the sample. Although conventional culture is considered as the gold standard for the diagnosis of bacterial LRTIs, it may be challenging to accurately recover all pathogens from a nonsterile sample. Alternatively, recent antibiotic treatment may affect the culture results and some target bacteria may present at a concentration <10^5^ CFU/mL and thus not detected by culture method. Mixed bacteria by PCRs with quantitative results above the culture positivity cut-off can be regarded as a recent or current mixed infection. On the other hand, mixed bacteria by PCRs with quantitative result(s) below the cut-off should be followed up by additional PCR testing to confirm the pathogens.

In this study, *S. pneumoniae* were only found by PCR assays in mixed infections, which was the main factor affecting the agreement between the two methods. *S. pneumoniae* was notoriously difficult to identify by conventional culture because the bacteria tends to autolyze after reaching the stationary phase, as well as the effect of prior antibiotic treatment ^21^. Previous studies comparing multiplex PCR with culture for the detection of *S. pneumoniae* and *H. influenzae* from sputum also found similar results with *S. pneumoniae* only found by qPCR ^22^. Our quantitative PCR assay therefore could serve as a useful diagnostic tool to detect *S. pneumoniae* from respiratory samples.

The use of EG-mPCR assays for AMR gene identification was also evaluated using 50 clinical samples. Overall, our findings demonstrated a high agreement between the detected AMR genes and the AMR profiles of target bacteria present in single infections. This means our assays can rapidly identify clinically relevant AMR genes to guide antibiotic therapy. As PCRs tend to detect more mixed infections, there may be some uncertainty about which AMR genes belong to which bacteria in these samples. However, the distribution of species-specific AMR genes in single infections and an understanding of local epidemiology of bacteria-drug resistance combinations can help to predict the most probable combinations of AMR gene-bacteria in mixed infections. Further research with increased sample size will provide more concrete data to discern AMR gene-bacteria combinations in mixed infections.

Our study has some limitations. The EG-mPCR assay had low sensitivity for the detection of *S. aureus*, probably due to its resistance to chemical lysis during DNA extraction ^23^. This could be improved in future investigation by sample treatment with lysozyme and lysostaphin enzyme during DNA extraction. Here we only looked for commonly acquired AMR genes in the target bacteria, disregarding the resistance mechanisms driven by single point mutations or overexpression of efflux pump, and thus the absence of AMR genes may not always correlate with a lack of phenotype. Furthermore, as we only target 6 key pathogens, the PCR-negative samples may contain other pathogens that are not included in our PCR assays.

In conclusion, our multiplex realtime-PCR with EvaGreen dye MCA assays have been proven to be sensitive and quantitative for rapid detection of 6 key pathogens and 14 AMR genes directly from respiratory samples. Our assays provide culture-independent information regarding bacterial pathogens and pathogen abundance in samples as well as the genotypic AMR status of LRTIs cases. Our method can possibly distinguish colonization and infection, offering an important tool to promote the optimal use of antibiotics and to monitor the clinical and antibiotic treatment response in patients with LRITs. The PCR assays have strong potentials to be adopted in clinical practice due to its feasibility, low cost and fast turnaround time. The potential impact of our PCR assays on antibiotic therapy warrants a thorough investigation to promote its implementation in clinical practice.

## Data Availability

All data produced in the present work are contained in the manuscript

## Acknowledgments

We would like to thank all members of the Molecular Epidemiology group at Oxford University Clinical Research Unit (OUCRU) in Vietnam and the Microbiology Laboratory at Hospital for Tropical Diseases for their support during the conduct of the study.

## Financial Support

Dr Pham Thanh Duy is funded by Wellcome International Training Fellowship (grant number: 222983/Z/21/Z). The funders had no role in study design, data collection and analysis, decision to publish, or preparation of the manuscript.

## Conflicts of interest

The authors declare no competing interests.

## Notes

### Competing Interest Statement

The authors have declared no competing interest.

### Funding Statement

This study was funded by Wellcome

